# Biological aging measures based on blood DNA methylation and risk of cancer: a prospective study

**DOI:** 10.1101/2020.04.08.20058727

**Authors:** Pierre-Antoine Dugué, Julie K Bassett, Ee Ming Wong, JiHoon E Joo, Shuai Li, Chenglong Yu, Daniel F Schmidt, Enes Makalic, Nicole Wong Doo, Daniel D Buchanan, Allison M Hodge, Dallas R English, John L Hopper, Graham G Giles, Melissa C Southey, Roger L Milne

## Abstract

**Background:** We previously investigated the association between five ‘first-generation’ measures of epigenetic aging and cancer risk in the Melbourne Collaborative Cohort Study. The present study assessed cancer risk associations for three recently developed methylation-based biomarkers of aging: *PhenoAge, GrimAge*, and predicted telomere length.

**Methods:** We estimated rate ratios (RRs) for the association between these three age-adjusted measures and risk of colorectal (N=813), gastric (N=165), kidney (N=139), lung (N=327), mature B-cell (N=423), prostate (N=846) and urothelial (N=404) cancer, using conditional logistic regression models. We also assessed associations by time since blood draw and by cancer subtype, and investigated potential non-linearity.

**Results:** We observed relatively strong associations of age-adjusted *PhenoAge* with risk of colorectal, kidney, lung, mature B-cell, and urothelial cancers (RR per standard deviation [SD]∼1.2-1.3). Similar findings were obtained for age-adjusted *GrimAge*, but the association with lung cancer risk was much larger, RR per SD=1.82, 95%CI=1.44-2.30, after adjustment for smoking status, pack-years, starting age, time since quitting and other cancer risk factors. Most associations appeared linear, larger than for the first-generation measures, and were virtually unchanged after adjustment for a large set of sociodemographic, lifestyle and anthropometric variables. For cancer overall, the comprehensively-adjusted RR per SD was 1.13, 95%CI=1.07-1.19, for *PhenoAge* and 1.12, 95%CI=1.05-1.20, for *GrimAge*, and appeared larger within 5 years of blood draw (RR=1.29 and 1.19, respectively).

**Conclusion:** The methylation-based measures *PhenoAge* and *GrimAge* may provide insights into the relationship between biological aging and cancer and be useful to predict cancer risk, particularly for lung cancer.

## INTRODUCTION

DNA methylation is one of the key mechanisms thought to underlie the association between aging and cancer **[1, 2]**. Biological aging measures derived from blood DNA methylation - taking advantage of varying rates of aging-associated methylation changes between individuals - have gained considerable popularity as tools to better understand and predict disease **[3-6]**. We previously investigated the association between five ‘first-generation’ measures of epigenetic aging **[7-9]** and the risk of seven cancer types using data from the Melbourne Collaborative Cohort Study (MCCS) **[10]**. The observed associations were relatively weak compared with those obtained for all-cause mortality **[9]**; cancer risk overall was increased by 4% to 9% per five-year increase in methylation ‘age acceleration’, although these estimates varied by cancer type.

Two novel methylation-based measures of biological aging, called *PhenoAge* **[11]** and *GrimAge* **[12]**, have been developed based on associations of DNA methylation with, for *PhenoAge*, age, mortality and clinical biomarkers, and for *GrimAge*, smoking pack-years and plasma concentrations of adrenomedullin, beta-2 microglobulin, cystatin C, growth differentiation factor 15, leptin, plasminogen activation inhibitor 1, and tissue inhibitor metalloproteinase 1. These new measures have proved to be more strongly associated with mortality **[11]** than the first-generation measures. Telomere length is another widely used biomarker of aging, which shows unclear associations with cancer risk **[2, 13, 14]**. A methylation-based predictor of telomere length was recently developed **[15]**.

In this study, we aimed to assess the association of the three aforementioned measures of biological aging, calculated using DNA methylation data from the Infinium HumanMethylation450 (HM450) assay, and the risk of seven cancer types: colorectal, gastric, kidney, lung, prostate and urothelial cancers, and mature B-cell neoplasms. We used a prospective design and 3,117 incident cancer cases and matched controls were included in the analysis.

## METHODS

### Study sample and blood collection

We used data collected from participants in the MCCS, a prospective study of 41,513 adult volunteers (24,469 women) aged between 27 and 76 years (99.3% aged 40–69 years) when recruited between 1990 and 1994 **[16]**. DNA samples were collected from peripheral blood drawn at the time of recruitment (1990-1994) or at the wave 2 follow-up visit (2003-2007). The DNA source was dried blood spots, peripheral blood mononuclear cells or buffy coats for 70%, 28% and 2% of participants, respectively (**Supplementary Material**).

Study participants provided informed consent in accordance with the Declaration of Helsinki and the study was approved by Cancer Council Victoria’s Human Research Ethics Committee.

### Cancer case-control studies nested in the MCCS

A series of case-control studies nested within the MCCS of colorectal (N=835 pairs), gastric (N=170), kidney (N=143), lung (N=332), prostate (N=869), and urothelial cancers (N=428) mature B-cell neoplasms (N=439) were conducted **[17-20]**. Cancer diagnoses were identified by linkage with the Victorian Cancer Registry and the Australian Cancer Database (Australian Institute of Health and Welfare). For each nested case–control study, controls were individually matched to incident cases (diagnosed after blood sample collection) on age using incidence density sampling (i.e. they had to be free of the cancer of interest up to the age at diagnosis of the corresponding case), sex, country of birth (Australia/New-Zealand, Southern Europe, Northern Europe), blood DNA source (dried blood spots, peripheral blood mononuclear cells or buffy coat) and collection period (baseline or wave 2, the latter applicable to 151 case-control pairs of the urothelial cancer study). Controls were also matched to cases on year of birth, except for the colorectal cancer study where controls were matched on year of baseline attendance. For the lung cancer study, controls were also matched on smoking history (never; former, quitting <10 years; former, quitting ≥10 years; current, smoking <15 cigarettes/day; current smoking ≥15 cigarettes/day) at the time of blood collection. For each study, matched cases and controls were placed next to each other, but allocated randomly, on the same slide.

### DNA extraction and bisulfite conversion, and DNA methylation data processing

Methods relating to DNA extraction and bisulfite conversion, and DNA methylation data processing have been described previously **[21]** and are detailed in the **Supplementary Material**.

### Methylation-based measures of biological aging

*PhenoAge, GrimAge* and methylation-predicted telomere length, and their respective age-adjusted measures (the residual from the regression of biological age on chronological age) were obtained using Horvath’s online calculator at https://dnamage.genetics.ucla.edu/new **[7, 11, 12]**.

### Statistical analysis

Pearson correlations of the three aging measures with each other and with age were calculated for participants selected as controls. We used conditional logistic regression to calculate odds ratios, which are estimates of the rate ratios [RRs] when incidence density sampling matching is used **[22]**, for the associations between age-adjusted biological aging measures, per standard deviation (1SD), and the risk of cancer. In Model 1, no covariates were included. In Model 2, we adjusted for smoking status (current / former / never), smoking pack-years, age at starting smoking (never smoked; age 16 or less; between age 17 and 21; after age 21 years); years since quitting smoking (never smoked; more than 10 years without smoking, between 5 and 10 years without smoking; less than 5 years without smoking), BMI (in kg/m2), height (in metres), alcohol intake in the past week (in grams/day), physical activity (categorised score based on time spent doing vigorous/less vigorous activities **[23]**), dietary quality (Alternative Healthy Eating Index 2010, **[24]**), socioeconomic status (deciles of the relative socioeconomic disadvantage of area of residence index **[25]**), education (ordinal variable ranging from 1: “primary school only” to 8:“tertiary or higher university degree”), **Table 1**. In Model 3, we added to Model 2 the white blood cell proportions estimated using the Houseman algorithm **[26]**. These models were used to analyse each cancer type separately, and all seven cancers combined; for the combined analysis, where an individual was diagnosed with several cancers, we included the first diagnosis only (respecting the incidence density sampling procedure), so that participants did not contribute twice to the pooled estimate. Analyses were additionally stratified (Model 1) by time between blood draw and diagnosis of the case (≤5, 5-10, and >10 years) and effect modification was examined using likelihood ratio tests of the interaction between each measure and the time-to-diagnosis variable, used as either categorical (P-heterogeneity) or continuous (P-linearity). Potential non-linearity in the associations between methylation-based measures and cancer risk was assessed using penalised regression splines, specifically P-splines, which are based on cubic B-splines and a large number of equidistant knots **[27]**, with three degrees of freedom. This type of spline was chosen because results are numerically stable, not sensitive to the location and number of knots **[28]**. These were represented graphically and non-linearity was assessed by comparing the P-spline and linear models using a likelihood ratio test. Case-control pairs with any missing values for the confounders (Model 2) measured at baseline were excluded and missing values at follow-up (urothelial cancers) were replaced by baseline values; 3% of the initial sample was excluded due to missing values. We also excluded 6 case-control pairs (0.2%) for which a participant had an outlying value (>5 or ≤ −5) for any of the three age-adjusted methylation-based measures (**Supplementary Figure 1**). The same models were used to calculate associations: i) expressed for a five-year increase, for *PhenoAge* and *GrimAge* (**Supplementary Table 4**); ii) expressed per a 1SD increase, for the first-generation measures (**Supplementary Table 5**).

**Table 1.** Characteristics of the study sample, seven case-control studies nested within the Melbourne Collaborative Cohort Study (N=41,513)

|  | Controls | Cases |
| --- | --- | --- |
| Cancer type (N) |  |  |
| Colorectal cancer | 813 | 813 |
| Gastric cancer | 165 | 165 |
| Kidney cancer | 139 | 139 |
| Lung cancer | 327 | 327 |
| Mature B-cell neoplasms | 423 | 423 |
| Prostate cancer | 846 | 846 |
| Urothelial cancers | 404 | 404 |
| Matching variables |  |  |
| Age at blood draw (years), median [IQR] | 61 [54-66] | 61 [54-66] |
| Sex: male, N (%) | 2,159 (69%) | 2,159 (69%) |
| Sex: female, N (%) | 958 (31%) | 958 (31%) |
| Country of birth: Australia/NZ, N (%) | 2,079 (67%) | 2,094 (67%) |
| Northern Europe, N (%) | 211 (7%) | 205 (7%) |
| Southern Europe, N (%) | 827 (26%) | 818 (26%) |
| Blood sample type: Dried blood spots, N (%) | 2,142 (69%) | 2,142 (69%) |
| Peripheral blood mononuclear cells, N (%) | 794 (25%) | 794 (25%) |
| Buffy coats, N (%) | 181 (6%) | 181 (6%) |
| Potential confounders |  |  |
| Smoking: current, N (%) | 458 (15%) | 485 (16%) |
| Smoking: former, N (%) | 1,230 (39%) | 1,294 (41%) |
| Smoking: never, N (%) | 1,429 (46%) | 1,338 (43%) |
| Smoking pack-years, median [IQR] | 2.4 [0-27.2] | 4.5 [0-31.1] |
| Age at starting: never smoked, N (%) | 1,429 (46%) | 1,338 (43%) |
| Age at starting: ≤16 years, N (%) | 639 (20%) | 672 (22%) |
| Age at starting: 17-21 years, N (%) | 749 (24%) | 834 (27%) |
| Age at starting: ≥22 years, N (%) | 300 (10%) | 273 (9%) |
| Time since quitting: never smoked, N (%) | 1,429 (46%) | 1,338 (43%) |
| Time since quitting: <5 years, N (%) | 622 (20%) | 675 (22%) |
| Time since quitting: 5-10 years, N (%) | 862 (28%) | 885 (28%) |
| Time since quitting: >10 years, N (%) | 204 (7%) | 219 (7%) |
| Body mass index (kg/m<sup>2</sup>), median [IQR] | 27 [24-29] | 27 [25-30] |
| Height (m), median [IQR] | 168 [161-174] | 169 [162-175] |
| Alcohol consumption (g/day), median [IQR] | 4 [0-19] | 5 [0-19.3] |
| Diet quality: AHEI-2010, median [IQR] | 63 [56-71] | 63 [56-71] |
| Physical activity score, median [IQR] | 2 [1-4] | 2 [1-4] |
| Education score, median [IQR] | 4 [4-6] | 4 [4-6] |
| Socioeconomic status: SEIFA-10, median [IQR] | 5 [3-8] | 6 [3-9] |

**Figure 1.**
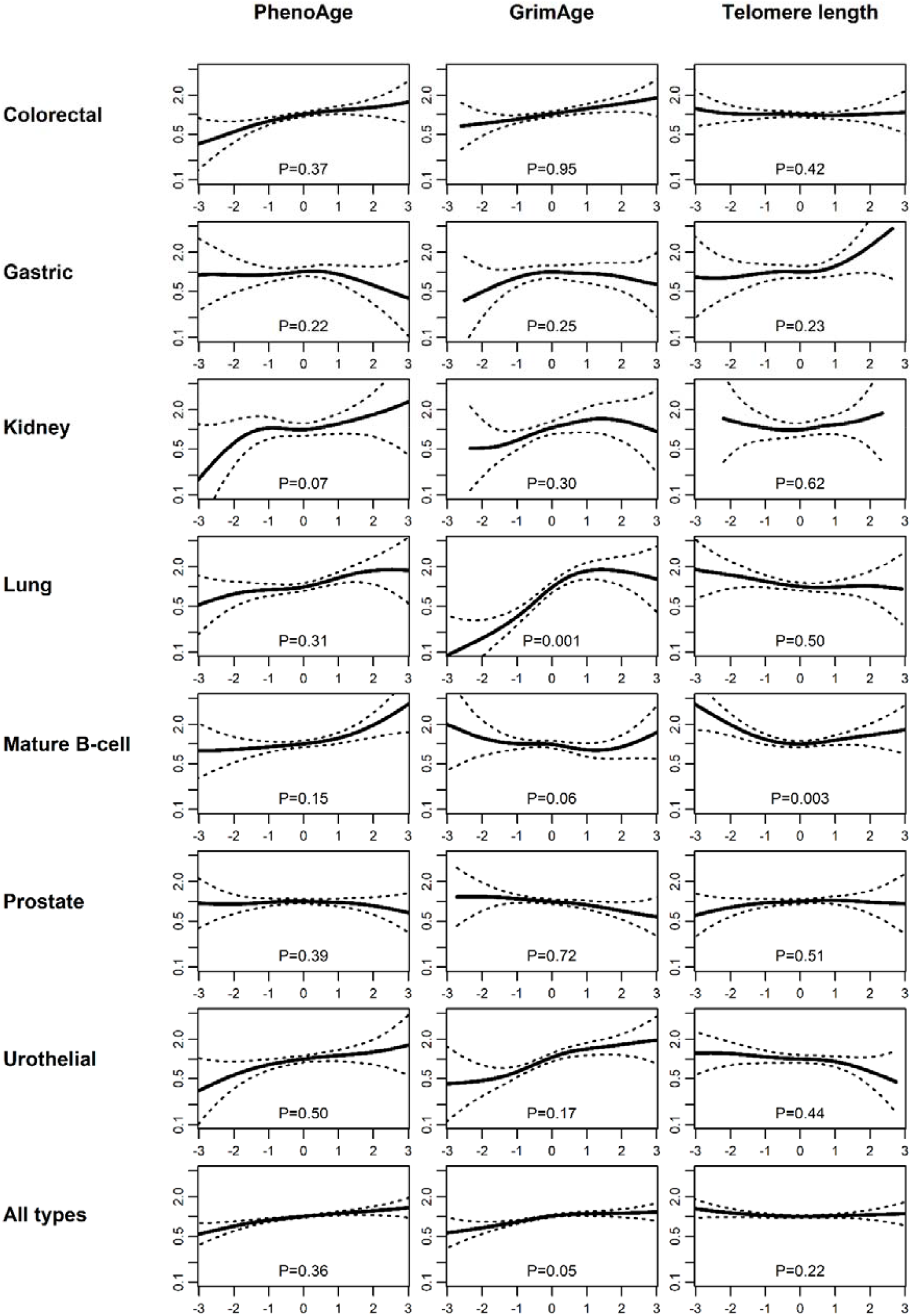
Relative cancer rates for age-adjusted *PhenoAge, GrimAge* and predicted telomere length for seven cancer types and overall in the Melbourne Collaborative Cohort Study. Legend: Model 1 was used (no other adjustment than that provided by the matching variables. X-axis: Methylation-based measures of aging; All measures were expressed as Z-scores (mean=0, standard deviation=1), so that ∼95% of the values are between – 2 and 2. Y-axis: Relative cancer rate, using as a reference (Y=1) the median value of the methylation-based measure distribution. P: P-value from a likelihood ratio test comparing P-spline and linear model.

In secondary analyses, we assessed the association between biological aging measures and risk of the following cancer subtypes, as defined in previous publications: colon / rectal cancer; multiple myeloma / follicular lymphoma / low-grade non-Hodgkin’s lymphoma (including chronic lymphocytic leukemia) / high-grade non-Hodgkin’s lymphoma **[20]**, aggressive / non-aggressive prostate cancer **[18]**, and invasive / superficial urothelial cancers **[19]**.

## RESULTS

The correlation with chronological age was 0.70, 0.80 and −0.55 for *PhenoAge, GrimAge* and methylation-predicted telomere length, respectively. For the age-adjusted measures, the correlation between *PhenoAge* and *GrimAge* was 0.34, and their correlations with methylation-predicted telomere length were −0.25 and −0.29, respectively (**Supplementary Table 1**). The correlations of the three measures with the five first-generation measures of epigenetic aging (all age-adjusted) were in the same range (**Supplementary Table 2)**.

**Table 2.**
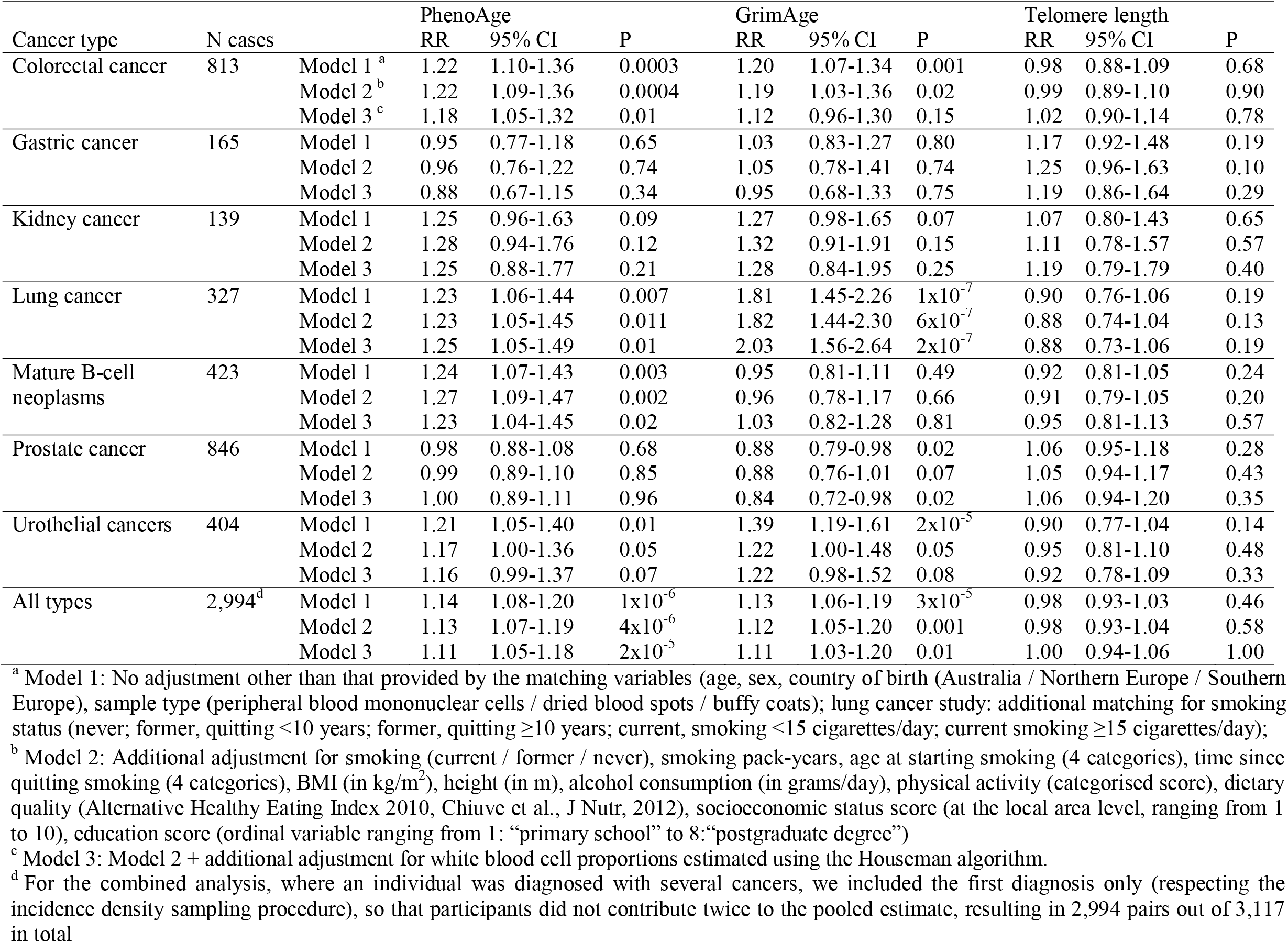
Association (Rate Ratios, 95% confidence intervals) between three methylation-based measures of aging (per one standard deviation) and cancer risk in the Melbourne Collaborative Cohort Study

| Cancer type | N cases |  | PhenoAge |  |  | GrimAge |  |  | Telomere length |  |  |
| --- | --- | --- | --- | --- | --- | --- | --- | --- | --- | --- | --- |
|  |  |  | RR | 95% CI | P | RR | 95% CI | P | RR | 95% CI | P |
| Colorectal cancer | 813 | Model 1<sup>a</sup> | 1.22 | 1.10-1.36 | 0.0003 | 1.20 | 1.07-1.34 | 0.001 | 0.98 | 0.88-1.09 | 0.68 |
|  |  | Model 2<sup>b</sup> | 1.22 | 1.09-1.36 | 0.0004 | 1.19 | 1.03-1.36 | 0.02 | 0.99 | 0.89-1.10 | 0.90 |
|  |  | Model 3<sup>c</sup> | 1.18 | 1.05-1.32 | 0.01 | 1.12 | 0.96-1.30 | 0.15 | 1.02 | 0.90-1.14 | 0.78 |
| Gastric cancer | 165 | Model 1 | 0.95 | 0.77-1.18 | 0.65 | 1.03 | 0.83-1.27 | 0.80 | 1.17 | 0.92-1.48 | 0.19 |
|  |  | Model 2 | 0.96 | 0.76-1.22 | 0.74 | 1.05 | 0.78-1.41 | 0.74 | 1.25 | 0.96-1.63 | 0.10 |
|  |  | Model 3 | 0.88 | 0.67-1.15 | 0.34 | 0.95 | 0.68-1.33 | 0.75 | 1.19 | 0.86-1.64 | 0.29 |
| Kidney cancer | 139 | Model 1 | 1.25 | 0.96-1.63 | 0.09 | 1.27 | 0.98-1.65 | 0.07 | 1.07 | 0.80-1.43 | 0.65 |
|  |  | Model 2 | 1.28 | 0.94-1.76 | 0.12 | 1.32 | 0.91-1.91 | 0.15 | 1.11 | 0.78-1.57 | 0.57 |
|  |  | Model 3 | 1.25 | 0.88-1.77 | 0.21 | 1.28 | 0.84-1.95 | 0.25 | 1.19 | 0.79-1.79 | 0.40 |
| Lung cancer | 327 | Model 1 | 1.23 | 1.06-1.44 | 0.007 | 1.81 | 1.45-2.26 | 1x10<sup>-7</sup> | 0.90 | 0.76-1.06 | 0.19 |
|  |  | Model 2 | 1.23 | 1.05-1.45 | 0.011 | 1.82 | 1.44-2.30 | 6x10<sup>-7</sup> | 0.88 | 0.74-1.04 | 0.13 |
|  |  | Model 3 | 1.25 | 1.05-1.49 | 0.01 | 2.03 | 1.56-2.64 | 2x10<sup>-7</sup> | 0.88 | 0.73-1.06 | 0.19 |
| Mature B-cell neoplasms | 423 | Model 1 | 1.24 | 1.07-1.43 | 0.003 | 0.95 | 0.81-1.11 | 0.49 | 0.92 | 0.81-1.05 | 0.24 |
|  |  | Model 2 | 1.27 | 1.09-1.47 | 0.002 | 0.96 | 0.78-1.17 | 0.66 | 0.91 | 0.79-1.05 | 0.20 |
|  |  | Model 3 | 1.23 | 1.04-1.45 | 0.02 | 1.03 | 0.82-1.28 | 0.81 | 0.95 | 0.81-1.13 | 0.57 |
| Prostate cancer | 846 | Model 1 | 0.98 | 0.88-1.08 | 0.68 | 0.88 | 0.79-0.98 | 0.02 | 1.06 | 0.95-1.18 | 0.28 |
|  |  | Model 2 | 0.99 | 0.89-1.10 | 0.85 | 0.88 | 0.76-1.01 | 0.07 | 1.05 | 0.94-1.17 | 0.43 |
|  |  | Model 3 | 1.00 | 0.89-1.11 | 0.96 | 0.84 | 0.72-0.98 | 0.02 | 1.06 | 0.94-1.20 | 0.35 |
| Urothelial cancers | 404 | Model 1 | 1.21 | 1.05-1.40 | 0.01 | 1.39 | 1.19-1.61 | 2x10<sup>-5</sup> | 0.90 | 0.77-1.04 | 0.14 |
|  |  | Model 2 | 1.17 | 1.00-1.36 | 0.05 | 1.22 | 1.00-1.48 | 0.05 | 0.95 | 0.81-1.10 | 0.48 |
|  |  | Model 3 | 1.16 | 0.99-1.37 | 0.07 | 1.22 | 0.98-1.52 | 0.08 | 0.92 | 0.78-1.09 | 0.33 |
| All types | 2,994<sup>d</sup> | Model 1 | 1.14 | 1.08-1.20 | 1x10<sup>-6</sup> | 1.13 | 1.06-1.19 | 3x10<sup>-5</sup> | 0.98 | 0.93-1.03 | 0.46 |
|  |  | Model 2 | 1.13 | 1.07-1.19 | 4x10<sup>-6</sup> | 1.12 | 1.05-1.20 | 0.001 | 0.98 | 0.93-1.04 | 0.58 |
|  |  | Model 3 | 1.11 | 1.05-1.18 | 2x10<sup>-5</sup> | 1.11 | 1.03-1.20 | 0.01 | 1.00 | 0.94-1.06 | 1.00 |

<sup>a</sup> Model 1: No adjustment other than that provided by the matching variables (age, sex, country of birth (Australia / Northern Europe / Southern Europe), sample type (peripheral blood mononuclear cells / dried blood spots / buffy coats); lung cancer study: additional matching for smoking status (never; former, quitting <10 years; former, quitting ≥10 years; current, smoking <15 cigarettes/day; current smoking ≥15 cigarettes/day);
<sup>b</sup> Model 2: Additional adjustment for smoking (current / former / never), smoking pack-years, age at starting smoking (4 categories), time since quitting smoking (4 categories), BMI (in kg/m<sup>2</sup>), height (in m), alcohol consumption (in grams/day), physical activity (categorised score), dietary quality (Alternative Healthy Eating Index 2010, Chiuve et al., J Nutr, 2012), socioeconomic status score (at the local area level, ranging from 1 to 10), education score (ordinal variable ranging from 1: “primary school” to 8: “postgraduate degree”)
<sup>c</sup> Model 3: Model 2 + additional adjustment for white blood cell proportions estimated using the Houseman algorithm.
<sup>d</sup> For the combined analysis, where an individual was diagnosed with several cancers, we included the first diagnosis only (respecting the incidence density sampling procedure), so that participants did not contribute twice to the pooled estimate, resulting in 2,994 pairs out of 3,117 in total

Hereafter, the age-adjusted measures are referred to as *PhenoAge, GrimAge* and telomere length. Their associations with cancer risk are presented in **Table 2**. In models without adjustment other than that provided by the matching variables, increasing *PhenoAge* was associated with increased risk of several types of cancer, including colorectal cancer (per 1SD RR=1.22, 95%CI: 1.10-1.36), kidney cancer (RR=1.25, 95%CI: 0.96-1.63), lung cancer (RR=1.23, 95%CI: 1.06-1.44), mature B-cell neoplasms (RR=1.24, 95%CI: 1.07-1.43), urothelial cancer (RR=1.21, 95%CI: 1.05-1.40), and cancer overall (RR=1.14, 95%CI: 1.08-1.20). These RRs were virtually the same after adjustment for a comprehensive set of cancer risk factors (cancer overall, RR=1.13, 95%CI: 1.07-1.19). *GrimAge* biological aging showed similar associations to *PhenoAge* for risk of colorectal, kidney cancer and cancer overall. The association with risk of lung cancer was much stronger (per 1SD RR=1.81, 95%CI: 1.45-2.26). A possible inverse association with risk of prostate cancer was also observed (RR=0.88, 95%CI: 0.79-0.98). These associations were virtually the same in comprehensively adjusted models (lung cancer RR=1.82, 95%CI=1.44-2.30), except for urothelial cancer for which estimates showed substantial attenuation, while remaining quite strong (RR=1.22, 95%CI=1.00-1.48). The RR also remained similar after additional adjustment for estimated white blood cell proportions for cancer overall (RR=1.11), being somewhat smaller for colorectal cancer risk (*PhenoAge:* RR=1.18; *GrimAge*: RR=1.12) but larger for lung cancer risk (*GrimAge*: RR=2.03), **Table 2**. We found no association between methylation-predicted telomere length and risk of any type of cancer or cancer overall (all P>0.1).

In analyses stratified by time since blood draw (**Table 3**), associations were somewhat larger within 5 years of blood draw for several cancer types for *PhenoAge:* colorectal cancer, RR=1.48 (P-linearity=0.07), lung cancer, RR=1.51 (P-linearity=0.18), and mature B-cell neoplasms, RR=1.38 (P-linearity=0.14), and this pattern was even clear in the overall cancer analysis (RR=1.29, 1.12 and 1.09 for ≤5, 5-10 and >10 years, respectively; P-linearity=0.004). A similar trend, albeit weaker, was observed for *GrimAge* (RR=1.19, 1.15, 1.08, respectively; P-linearity=0.11). As shown in **Figure 1**, most associations with cancer risk appeared relatively linear. Some evidence of non-linearity was observed for *GrimAge* and lung cancer risk (P=0.001), with a sharp increase at lower values and a plateau after the 75^th^ percentile. A similar shape of association, while less marked, was also observed for *GrimAge* and overall cancer risk (P=0.05).

**Table 3.** Stratification by time since blood draw for the association (Rate Ratios, 95% confidence intervals^a^) between three methylation-based measures of aging (per one standard deviation) and cancer risk in the Melbourne Collaborative Cohort Study

| Cancer type | N cases | Time since blood draw | PhenoAge |  |  |  | GrimAge |  |  |  | Telomere length |  |  |  |
| --- | --- | --- | --- | --- | --- | --- | --- | --- | --- | --- | --- | --- | --- | --- |
|  |  |  | RR<sup>a</sup> | 95% CI | P-het<sup>b</sup> | P-lin<sup>b</sup> | RR<sup>a</sup> | 95% CI | P-het<sup>b</sup> | P-lin<sup>b</sup> | RR<sup>a</sup> | 95% CI | P-het<sup>b</sup> | P-lin<sup>b</sup> |
| Colorectal cancer | 813 | ≤ 5 years | 1.48 | 1.16-1.89 |  |  | 1.20 | 0.94-1.52 |  |  | 0.95 | 0.75-1.20 |  |  |
|  |  | 5-10 years | 1.23 | 1.02-1.50 | 0.13 | 0.07 | 1.24 | 1.03-1.50 | 0.89 | 0.32 | 1.05 | 0.87-1.26 | 0.68 | 0.79 |
|  |  | > 10 years | 1.11 | 0.95-1.30 |  |  | 1.17 | 0.99-1.38 |  |  | 0.95 | 0.81-1.10 |  |  |
| Gastric cancer | 165 | ≤ 5 years | 0.72 | 0.42-1.26 |  |  | 0.99 | 0.54-1.84 |  |  | 0.98 | 0.57-1.71 |  |  |
|  |  | 5-10 years | 1.06 | 0.71-1.59 | 0.52 | 0.48 | 0.93 | 0.61-1.42 | 0.85 | 0.49 | 1.14 | 0.73-1.77 | 0.74 | 0.60 |
|  |  | > 10 years | 0.97 | 0.73-1.30 |  |  | 1.08 | 0.82-1.41 |  |  | 1.26 | 0.91-1.74 |  |  |
| Kidney cancer | 139 | ≤ 5 years | 1.29 | 0.61-2.70 |  |  | 0.81 | 0.46-1.42 |  |  | 1.68 | 0.82-3.45 |  |  |
|  |  | 5-10 years | 1.33 | 0.72-2.48 | 0.97 | 0.67 | 1.91 | 1.06-3.44 | 0.09 | 0.26 | 1.01 | 0.54-1.91 | 0.36 | 0.13 |
|  |  | > 10 years | 1.22 | 0.89-1.67 |  |  | 1.29 | 0.89-1.87 |  |  | 0.95 | 0.66-1.38 |  |  |
| Lung cancer | 327 | ≤ 5 years | 1.51 | 1.05-2.18 |  |  | 2.15 | 1.28-3.61 |  |  | 0.76 | 0.51-1.14 |  |  |
|  |  | 5-10 years | 1.13 | 0.85-1.50 | 0.44 | 0.18 | 1.46 | 1.00-2.12 | 0.40 | 0.92 | 0.97 | 0.71-1.31 | 0.64 | 0.51 |
|  |  | > 10 years | 1.20 | 0.97-1.49 |  |  | 1.94 | 1.40-2.69 |  |  | 0.91 | 0.73-1.13 |  |  |
| Mature B-cell neoplasms | 423 | ≤ 5 years | 1.38 | 1.01-1.90 |  |  | 0.85 | 0.58-1.25 |  |  | 0.94 | 0.72-1.22 |  |  |
|  |  | 5-10 years | 1.21 | 0.93-1.57 | 0.73 | 0.14 | 1.22 | 0.87-1.71 | 0.22 | 0.30 | 1.03 | 0.78-1.35 | 0.64 | 0.30 |
|  |  | > 10 years | 1.20 | 0.98-1.46 |  |  | 0.88 | 0.71-1.09 |  |  | 0.88 | 0.73-1.05 |  |  |
| Prostate cancer | 846 | ≤ 5 years | 1.23 | 0.96-1.57 |  |  | 0.90 | 0.71-1.13 |  |  | 1.03 | 0.82-1.30 |  |  |
|  |  | 5-10 years | 0.93 | 0.77-1.13 | 0.13 | 0.13 | 0.94 | 0.74-1.18 | 0.78 | 0.95 | 1.09 | 0.88-1.34 | 0.95 | 0.95 |
|  |  | > 10 years | 0.93 | 0.81-1.07 |  |  | 0.85 | 0.73-0.99 |  |  | 1.06 | 0.91-1.23 |  |  |
| Urothelial cancers | 404 | ≤ 5 years | 1.17 | 0.94-1.47 |  |  | 1.70 | 1.31-2.22 |  |  | 0.90 | 0.72-1.13 |  |  |
|  |  | 5-10 years | 1.16 | 0.89-1.51 | 0.76 | 0.93 | 1.07 | 0.84-1.38 | 0.04 | 0.52 | 1.04 | 0.80-1.36 | 0.22 | 0.47 |
|  |  | > 10 years | 1.32 | 0.99-1.75 |  |  | 1.48 | 1.10-2.00 |  |  | 0.74 | 0.54-1.00 |  |  |
| All types | 2,994 | ≤ 5 years | 1.29 | 1.15-1.44 |  |  | 1.19 | 1.06-1.33 |  |  | 0.95 | 0.85-1.06 |  |  |
|  |  | 5-10 years | 1.12 | 1.01-1.23 | 0.05 | 0.004 | 1.15 | 1.04-1.28 | 0.36 | 0.11 | 1.05 | 0.95-1.16 | 0.31 | 0.70 |
|  |  | > 10 years | 1.09 | 1.01-1.17 |  |  | 1.08 | 1.00-1.17 |  |  | 0.96 | 0.89-1.03 |  |  |

<sup>a</sup> Model 1: No adjustment other than that provided by the matching variables (age, sex, country of birth (Australia / Northern Europe / Southern Europe), sample type (peripheral blood mononuclear cells / dried blood spots / buffy coats); lung cancer study: additional matching for smoking status (never; former, quitting <10 years; former, quitting ≥10 years; current, smoking <15 cigarettes/day; current smoking ≥15 cigarettes/day)
<sup>b</sup> P-het (P value for heterogeneity) and P-lin (P-value for linearity) were calculated using a likelihood ratio test for the interaction between each methylation-based measure and the time-to-diagnosis variable, taken as categorical and continuous, respectively.

Associations were generally consistent across cancer subtypes (**Supplementary Table 3**). Evidence of heterogeneity was observed for the association of *PhenoAge* with B-cell lymphoma subtypes (P=0.05, being stronger for low-grade non-Hodgkin lymphoma: RR=1.90, 95%CI: 1.37-2.62). Weak evidence of heterogeneity was observed for *PhenoAge* and colorectal cancer risk (P=0.16; colon: RR=1.16, rectum: RR=1.38). The inverse association observed between *GrimAge* and prostate cancer risk was only apparent for non-aggressive disease, RR=0.79 (95%CI: 0.64-0.97), P-heterogeneity=0.16. No association was found between methylation-predicted telomere length and risk of cancer subtypes.

## DISCUSSION

In this prospective study, including a total of 3,117 incident cancer cases, we observed relatively strong associations of *PhenoAge* and *GrimAge* with risk of several cancer types; these appeared to be greater than in our study of first-generation epigenetic aging measures for risk of colorectal, lung and urothelial cancer (**Supplementary Table 5**) **[10]**. For *GrimAge*, a very strong association was observed with risk of lung cancer, independently of several questionnaire-collected variables relating to smoking. An association stronger than with *PhenoAge* was also observed with risk of urothelial cancer. A possible inverse association was observed between *GrimAge* and (non-aggressive) prostate cancer. No association was observed between methylation-predicted telomere length and any cancer type or subtype.

*PhenoAge* and *GrimAge* integrate methylation measures at CpG sites associated with age, mortality, key disease risk factors and biomarkers, which are also involved in the aetiology of cancer. That *GrimAge* is enriched for smoking-associated methylation measures likely explains the very strong association observed with lung cancer risk; of note, however, case-control pairs were matched on smoking history in the lung cancer study and the estimates were robust to further adjustment for questionnaire-collected variables. In the case of urothelial cancer, for which smoking is a strong risk factor, the association was partially attenuated after adjustment for smoking status, which was not a matching variable in that study. However, for both *PhenoAge* and *GrimAge*, there was overall little attenuation of risk estimates after adjustment for a comprehensive set of sociodemographic and lifestyle cancer risk factors, which may indicate that these measures capture information beyond self-reported questionnaires and on many adverse environmental / lifestyle factors that affect the methylome over the lifecourse. To our knowledge, no data exist on the association of *PhenoAge* and *GrimAge* with risk of cancers other than pancreatic cancer **[29]**, and breast cancer, in the latter case for which the Sister Study revealed a reasonably strong association with *PhenoAge* **[30]** (invasive disease; hazard ratio per 5-year: 1.13, which is of similar magnitude to our findings for colorectal, kidney, lung, mature B-cell and urothelial cancers, **Supplementary Table 4**) but not with *GrimAge* **[31]**. Further adjustment for estimated white blood cell proportions slightly attenuated associations with cancer risk overall, although a larger association was observed for *GrimAge* and lung cancer, similar to observations made for *PhenoAge* and risk of breast **[30]** and pancreatic cancer **[29]**.

Although our sample size was quite large, our findings should be replicated by other studies before these methylation-based measures can be used for cancer risk prediction. In the case of lung cancer, our RR estimate of 1.8 per standard deviation for *GrimAge* is considerably larger than current estimates obtained for polygenic risk scores **[32-35]**. For other cancers, our estimates are lower than for polygenic risk scores for colorectal, gastric, B-cell lymphoma, and prostate cancer, and similar or greater for kidney and urothelial cancer **[32-35]**. Combining polygenic and methylation aging scores may therefore be required to summarise risk associated with genetic factors and lifestyle/environmental exposures accumulated over the lifetime. Our findings also suggest that *PhenoAge* and *GrimAge* may be more valuable biomarkers than the first-generation aging clocks **[10, 30]** and generally show a linear association with risk. That we observed stronger associations within five years of blood draw for *PhenoAge*, and to a lesser extent for *GrimAge*, suggests that these aging measures may have more utility for assessment of short-term risk, but corroborating data are required to confirm this. In our previous report on the first-generation measures **[10]**, we found at best weak evidence of effect modification by time since blood draw. Consistent with this, it was observed that Horvath epigenetic aging was largely determined before adulthood **[36]**, and this might not hold true for *PhenoAge* and *GrimAge* since these predictors were developed to predict a composite phenotype (age and clinical markers). Finally, while these methylation-based predictors show some degree of correlation with age in other tissues **[11]**, they were developed and validated in blood, so at this stage should be considered as biomarkers of future cancer risk and extrapolation to cancer or normal-adjacent tissue made with caution **[37]**, because DNA methylation usually shows substantial variation across tissues **[38]**.

We also used DNA methylation measures at a set of 140 CpGs to estimate telomere length. The correlation of this predictor with measured telomere length in independent data has been shown to be moderate (r ∼ 0.40), but its correlation with age appeared stronger than was the case for measured telomere length (r ∼ −0.75 vs −0.35), which is consistent with our findings (correlation with age r = −0.56). Our findings of no association between telomere length and cancer risk are consistent with those reported in a Danish prospective study of 3,142 cancer cases of any type **[14]**. In a Mendelian randomization study and meta-analysis by Haycock et al., which included a larger number of cancer cases and types, genetically-predicted telomere length was strongly positively associated with risks of lung and bladder cancers, which is inconsistent with our findings. Our results were nevertheless consistent with Mendelian randomization estimates for other cancer types and with estimates from prospective studies for all cancer types, all showing null or weak associations with cancer risk **[13, 14]**.

We conclude that biological aging, as defined by the methylation-based measures *PhenoAge* and *GrimAge*, is associated with risk of several cancer types, including colorectal, lung, kidney and urothelial cancers, and mature B-cell neoplasms, independently of key demographic, lifestyle and socioeconomic variables. These measures, derived using a limited number of methylation sites across the genome, have the potential to improve cancer risk prediction, particularly in contexts where relevant cancer biomarkers have not been extensively measured.

## Data Availability

The data that support the findings of this study are available from the corresponding author, PAD, upon reasonable request.

## NOTES

### Funding

This work was supported by the Australian National Health and Medical Research Council (NHMRC) grants 1164455. MCCS cohort recruitment was funded by VicHealth and Cancer Council Victoria. The MCCS was further supported by Australian NHMRC grants 209057, 251553 and 504711 and by infrastructure provided by Cancer Council Victoria. The nested case-control methylation studies were supported by the NHMRC grants 1011618, 1026892, 1027505, 1050198, 1043616 and 1074383. S.L. is a Victorian Cancer Agency Early Career Research Fellow (ECRF19020). M.C.S. is an NHMRC Senior Research Fellow (1061177). This work also received funding from Monash University, Melbourne, Australia.

### Role of the Funder

The funder played no part in the design, analysis, or drafting of this manuscript.

## Acknowledgements

Cases were ascertained through the Victorian Cancer Registry (VCR) and the Australian Cancer Database (Australian Institute of Health and Welfare).

## Data Availability Statement

The data that support the findings of this study are available from the corresponding author upon reasonable request.

## Author Contributions

Conceptualisation: all authors; data collection and curation: JKB, EMW and JEJ; formal analysis and visualisation: PAD; methodology: all authors; funding and resources: DS, EM, NWD, DDB, AMH, DRE, GGG, MCS, and RLM, writing – original draft: PAD; writing – review & editing: all authors.

## Conflicts of interest

None declared.

## Notes

### Competing Interest Statement

The authors have declared no competing interest.

### Summary of Updates

- changed from a brief report to a full article - several additional analyses: associations by time since blood draw, non-linearity of associations, and additional adjustments - augmented Discussion section

