## Supplementary Methods and Tables for "Biological aging measures based on blood DNA methylation and risk of cancer: a prospective study"

**DNA extraction and bisulfite conversion, and DNA methylation data processing**

*Bisulphite conversion of DNA and Illumina Infinium methylation assay*

Genomic DNA was extracted from mononuclear cells using the QIAamp 96 DNA blood kit (Qiagen), and from Guthrie card samples as previously described **[1].** Briefly, twenty blood spots of 3.2 mm diameter were punched from the Guthrie card and lysed in phosphate buffered saline using TissueLyser (Qiagen). The resulting supernatant was processed using Qiagen mini spin columns according to the manufacturer’s protocol. The quality and quantity of DNA was assessed using the Quant-iT™ Picogreen® dsDNA assay measured on the Qubit® Fluorometer (Life Technologies, Grand Island, NY), with a minimum of 0.3 μg DNA considered acceptable for methylation analysis. Genomic DNA was bisulphite converted using the EZ DNA Methylation-Gold kit (Zymo Research, Irvine, CA) following the manufacturer’s instructions. The Illumina Infinium HumanMethylation450K BeadChip (HM450K) array (Illumina, Inc.; San Diego, CA, USA) was used to measure DNA methylation. This array covers 99% of the RefSeq genes and uses a combination of two distinct probe types (Infinium I and II) to detect the methylation status of 485,577 CpGs in the human genome at a single base resolution **[2].** Samples in each nested case-control study and the longitudinal study were assayed at non-overlapping time periods. In each study, samples were randomly assigned to chips and processed as per the Illumina protocol. All laboratory work, including DNA extraction, bisulphite conversion and Infinium assaying was performed at the Genetic Epidemiology Laboratory, University of Melbourne.

***Normalization and quality control of methylation data***

The same normalization and quality control procedures were applied to methylation data in all eight studies (case-control and longitudinal). Raw .IDAT files were imported into R using the *minfi* package **[3].** Illumina’s background correction was applied based on internal control probes. Subset-quantile within-array normalisation (SWAN) was used to correct for the technical variability between the two probe types of the HM450K array **[4]**. The ‘getSex’ function of the *minfi* package **[3]** was used to predict sex for each sample. Samples for which predicted sex was inconsistent with recorded sex were excluded from further analyses. For each sample, each CpG with a detection *P* value >0.01 was assigned as missing. Samples with missing values for more than 5% of probes were excluded. CpG sites were excluded if they were missing for more than 20% of samples. β-values, which range from zero to one and correspond to the percentage of methylation, were calculated for each CpG using *minfi*. β-values were transformed into M-values using the formula: $M= {log}_{2}(\frac{\beta}{1-\beta})$. **[5]**.

**Supplementary Figure 1.** Exclusion of outliers based on the distribution of age-adjusted measures of biological aging (all participants), using 5 standard deviations as cut-off. Age-adjusted measures have mean=0 and variance=1. Six participants were identified as outliers and the corresponding case-control pairs were excluded
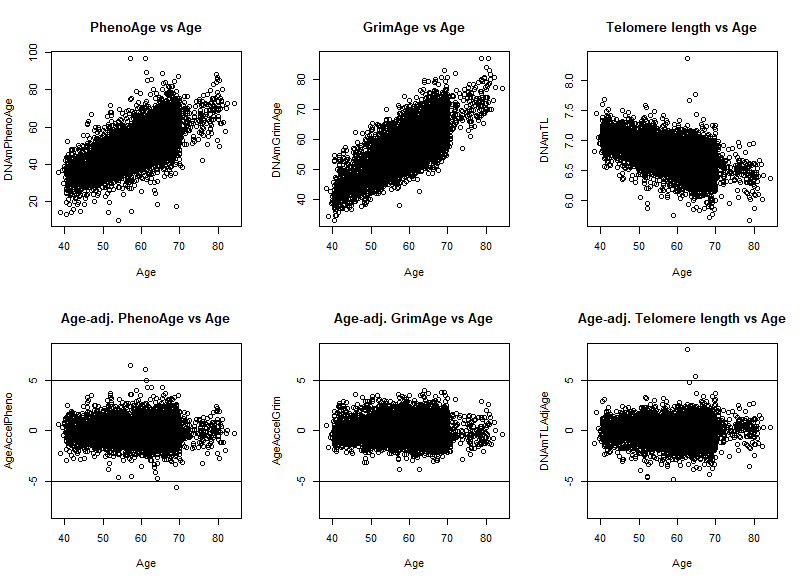

**Supplementary Table 1.** Pearson correlations between methylation-based predictors and chronological age

|  | **Chronological age** | **PhenoAge** | **GrimAge** | **Telomere length** | **Age-adjusted PhenoAge** | **Age-adjusted GrimAge** |
| --- | --- | --- | --- | --- | --- | --- |
| **PhenoAge** | 0.70 |  |  |  |  |  |
| **GrimAge** | 0.80 | 0.71 |  |  |  |  |
| **Telomere length** | -0.55 | -0.54 | -0.60 |  |  |  |
| **Age-adjusted PhenoAge** | 0.00 | 0.71 | 0.19 | -0.21 |  |  |
| **Age-adjusted GrimAge** | 0.00 | 0.24 | 0.57 | -0.24 | 0.34 |  |
| **Age-adjusted telomere length** | 0.00 | -0.18 | -0.17 | 0.83 | -0.25 | -0.29 |

**Supplementary Table 2.** Pearson correlations between three age-adjusted methylation measures of biological aging included in this study, and five age-adjusted first-generation measures presented in [6]

|  | **AA-Horvath** | **AA-Hannum** | **IEAA-Horvath** | **IEAA-Hannum** | **EEAA** |
| --- | --- | --- | --- | --- | --- |
| **Age-adjusted PhenoAge** | 0.35 | 0.39 | 0.35 | 0.29 | 0.41 |
| **Age-adjusted GrimAge** | 0.12 | 0.26 | 0.13 | 0.19 | 0.28 |
| **Age-adjusted telomere length** | -0.25 | -0.42 | -0.11 | -0.22 | -0.50 |

AA-Horvath: Age acceleration from the Horvath clock; AA-Hannum; Age acceleration from the Hannum clock; IEAA-Horvath: intrinsic epigenetic age acceleration from the Horvath clock; IEAA-Hannum: intrinsic epigenetic age acceleration from the Horvath clock; EEAA: enhanced Hannum age acceleration

**Supplementary Table 3.** Association (Rate Ratios (RR), 95% confidence intervals (95% CI) between three methylation-based measures of aging and cancer risk in the Melbourne Collaborative Cohort Study, by cancer subtypes.

| **Cancer type** | **Aging measure** |  |  | **RR** | **95% CI** | **P** | **Heterogeneity P (Model 2)** | |
| --- | --- | --- | --- | --- | --- | --- | --- | --- |
| **Colorectal cancer** |  |  |  |  |  |  |  |  |
|  | PhenoAge | Model 1 |  | 1.17 | 1.03-1.33 | 0.018 |  |  |
| **Colon (N=525)** |  | Model 2 |  | 1.16 | 1.01-1.32 | 0.029 |  |  |
|  | GrimAge | Model 1 |  | 1.15 | 1.00-1.31 | 0.043 |  |  |
|  |  | Model 2 |  | 1.16 | 0.98-1.38 | 0.088 |  | **P** |
|  | Telomere length | Model 1 |  | 0.99 | 0.86-1.12 | 0.832 | PhenoAge | 0.16 |
|  |  | Model 2 |  | 1.01 | 0.88-1.16 | 0.863 | GrimAge | 0.51 |
|  | PhenoAge | Model 1 |  | 1.34 | 1.11-1.62 | 0.003 | Telomere length | 0.74 |
| **Rectum (N=288)** |  | Model 2 |  | 1.38 | 1.12-1.69 | 0.003 |  |  |
|  | GrimAge | Model 1 |  | 1.32 | 1.08-1.60 | 0.006 |  |  |
|  |  | Model 2 |  | 1.29 | 1.00-1.66 | 0.048 |  |  |
|  | Telomere length | Model 1 |  | 0.97 | 0.82-1.15 | 0.695 |  |  |
|  |  | Model 2 |  | 0.97 | 0.81-1.17 | 0.779 |  |  |
| **B-cell mature neoplasm** |  |  |  |  |  |  |  |  |
|  | PhenoAge | Model 1 |  | 1.02 | 0.76-1.36 | 0.912 |  |  |
| **Multiple myeloma** |  | Model 2 |  | 1.15 | 0.82-1.60 | 0.425 |  |  |
| **(N=108)** | GrimAge | Model 1 |  | 1.02 | 0.74-1.41 | 0.901 |  |  |
|  |  | Model 2 |  | 0.96 | 0.62-1.50 | 0.870 |  |  |
|  | Telomere length | Model 1 |  | 0.88 | 0.67-1.15 | 0.356 |  |  |
|  |  | Model 2 |  | 0.84 | 0.62-1.14 | 0.254 |  |  |
|  | PhenoAge | Model 1 |  | 1.22 | 0.87-1.72 | 0.243 |  |  |
| **Follicular lymphoma** |  | Model 2 |  | 1.32 | 0.87-2.00 | 0.187 |  |  |
| **(N=78)** | GrimAge | Model 1 |  | 1.02 | 0.68-1.53 | 0.924 |  |  |
|  |  | Model 2 |  | 1.11 | 0.63-1.93 | 0.722 |  | **P** |
|  | Telomere length | Model 1 |  | 0.73 | 0.53-1.02 | 0.069 | PhenoAge | 0.05 |
|  |  | Model 2 |  | 0.72 | 0.48-1.08 | 0.116 | GrimAge | 0.52 |
|  | PhenoAge | Model 1 |  | 1.72 | 1.28-2.29 | 0.000 | Telomere length | 0.16 |
| **Low-grade NHL** |  | Model 2 |  | 1.90 | 1.37-2.62 | 0.000 |  |  |
| **(N=131)** | GrimAge | Model 1 |  | 0.98 | 0.75-1.27 | 0.865 |  |  |
|  |  | Model 2 |  | 1.14 | 0.79-1.63 | 0.485 |  |  |
|  | Telomere length | Model 1 |  | 1.15 | 0.91-1.46 | 0.235 |  |  |
|  |  | Model 2 |  | 1.15 | 0.88-1.49 | 0.312 |  |  |
|  | PhenoAge | Model 1 |  | 1.02 | 0.78-1.34 | 0.876 |  |  |
| **High-grade NHL (N=106)** |  | Model 2 |  | 1.08 | 0.80-1.45 | 0.607 |  |  |
|  | GrimAge | Model 1 |  | 0.77 | 0.55-1.10 | 0.152 |  |  |
|  |  | Model 2 |  | 0.76 | 0.49-1.17 | 0.208 |  |  |
|  | Telomere length | Model 1 |  | 0.85 | 0.66-1.10 | 0.230 |  |  |
|  |  | Model 2 |  | 0.82 | 0.62-1.08 | 0.159 |  |  |
| **Prostate cancer** |  |  |  |  |  |  |  |  |
|  | PhenoAge | Model 1 |  | 1.03 | 0.90-1.19 | 0.806 |  |  |
| **Prostate: aggressive** |  | Model 2 |  | 1.04 | 0.89-1.20 | 0.815 |  |  |
| **(N=417)** | GrimAge | Model 1 |  | 0.95 | 0.81-1.10 | 0.458 |  |  |
|  |  | Model 2 |  | 0.97 | 0.79-1.17 | 0.702 |  | **P** |
|  | Telomere length | Model 1 |  | 1.04 | 0.89-1.22 | 0.499 | PhenoAge | 0.35 |
|  |  | Model 2 |  | 1.04 | 0.89-1.23 | 0.494 | GrimAge | 0.16 |
|  | PhenoAge | Model 1 |  | 0.92 | 0.80-1.07 | 0.287 | Telomere length | 0.91 |
| **Prostate: non-aggressive** |  | Model 2 |  | 0.94 | 0.80-1.09 | 0.382 |  |  |
| **(N=429)** | GrimAge | Model 1 |  | 0.81 | 0.69-0.95 | 0.012 |  |  |
|  |  | Model 2 |  | 0.79 | 0.64-0.97 | 0.026 |  |  |
|  | Telomere length | Model 1 |  | 1.08 | 0.93-1.26 | 0.310 |  |  |
|  |  | Model 2 |  | 1.06 | 0.90-1.24 | 0.481 |  |  |
| **Urothelial cancer** |  |  |  |  |  |  |  |  |
|  | PhenoAge | Model 1 |  | 1.21 | 0.97-1.50 | 0.087 |  |  |
| **Urothelial: invasive** |  | Model 2 |  | 1.14 | 0.90-1.45 | 0.275 |  |  |
| **(N=182)** | GrimAge | Model 1 |  | 1.50 | 1.19-1.89 | 0.001 |  |  |
|  |  | Model 2 |  | 1.15 | 0.85-1.57 | 0.361 |  |  |
|  | Telomere length | Model 1 |  | 0.87 | 0.70-1.08 | 0.205 |  | **P** |
|  |  | Model 2 |  | 0.94 | 0.74-1.20 | 0.626 | PhenoAge | 0.83 |
|  | PhenoAge | Model 1 |  | 1.21 | 0.99-1.48 | 0.057 | GrimAge | 0.60 |
| **Urothelial: superficial** |  | Model 2 |  | 1.18 | 0.96-1.46 | 0.120 | Telomere length | 0.94 |
| **(N=222)** | GrimAge | Model 1 |  | 1.30 | 1.07-1.59 | 0.010 |  |  |
|  |  | Model 2 |  | 1.29 | 0.99-1.68 | 0.063 |  |  |
|  | Telomere length | Model 1 |  | 0.92 | 0.76-1.12 | 0.393 |  |  |
|  |  | Model 2 |  | 0.95 | 0.77-1.18 | 0.667 |  |  |

**Supplementary Table 4**. Association (Rate Ratios (RR), 95% confidence intervals (95% CI) between *PhenoAge* and *GrimAge* (per 5 years) and cancer risk in the Melbourne Collaborative Cohort Study

|  |  | |  | ***PhenoAge*** | | |  | ***GrimAge*** | | |
| --- | --- | --- | --- | --- | --- | --- | --- | --- | --- | --- |
| **Cancer type** | **N cases** | |  | **RR** | **95% CI** | **P** |  | **RR** | **95% CI** | **P** |
| **Colorectal cancer** | | 814 | Model 1 | 1.13 | 1.06-1.21 | 0.0005 |  | 1.22 | 1.08-1.39 | 0.001 |
|  |  |  | Model 2 | 1.13 | 1.05-1.21 | 0.0008 |  | 1.21 | 1.04-1.42 | 0.02 |
| **Gastric cancer** | | 166 | Model 1 | 0.96 | 0.84-1.11 | 0.60 |  | 1.03 | 0.81-1.30 | 0.84 |
|  |  |  | Model 2 | 0.97 | 0.83-1.13 | 0.69 |  | 1.04 | 0.75-1.45 | 0.81 |
| **Kidney cancer** | | 139 | Model 1 | 1.16 | 0.98-1.38 | 0.09 |  | 1.31 | 0.97-1.76 | 0.07 |
|  |  |  | Model 2 | 1.18 | 0.96-1.45 | 0.12 |  | 1.36 | 0.89-2.08 | 0.15 |
| **Lung cancer** | | 327 | Model 1 | 1.15 | 1.04-1.27 | 0.007 |  | 1.95 | 1.52-2.51 | 1x10^-7^ |
|  |  |  | Model 2 | 1.15 | 1.03-1.28 | 0.011 |  | 1.97 | 1.51-2.56 | 6x10^-7^ |
| **Mature B-cell neoplasms** | | 426 | Model 1 | 1.14 | 1.05-1.25 | 0.003 |  | 0.93 | 0.78-1.11 | 0.45 |
|  |  |  | Model 2 | 1.16 | 1.05-1.27 | 0.002 |  | 0.94 | 0.75-1.17 | 0.56 |
| **Prostate cancer** | | 847 | Model 1 | 0.98 | 0.92-1.05 | 0.58 |  | 0.86 | 0.76-0.98 | 0.02 |
|  |  |  | Model 2 | 0.99 | 0.92-1.06 | 0.74 |  | 0.86 | 0.74-1.01 | 0.07 |
| **Urothelial cancers** | | 404 | Model 1 | 1.13 | 1.03-1.25 | 0.01 |  | 1.45 | 1.22-1.72 | 2x10^-5^ |
|  |  |  | Model 2 | 1.11 | 1.00-1.22 | 0.05 |  | 1.25 | 1.00-1.55 | 0.05 |
| **All types** | | 3,086 | Model 1 | 1.09 | 1.05-1.12 | 3x10^-6^ |  | 1.14 | 1.07-1.21 | 5x10^-5^ |
|  |  |  | Model 2 | 1.08 | 1.04-1.12 | 9x10^-6^ |  | 1.13 | 1.05-1.22 | 0.002 |

**Model 1**: No adjustment other than that provided by the matching variables (age, sex, country of birth (Australia / Northern Europe / Southern Europe), sample type (peripheral blood mononuclear cells / dried blood spots / buffy coats); lung cancer study: additional matching for smoking status (never; former, quitting <10 years; former, quitting ≥10 years; current, smoking <15 cigarettes/day; current smoking ≥15 cigarettes/day)

**Model 2**: Additional adjustment for smoking (current / former / never), smoking pack-years, age at starting smoking (4 categories), time since quitting smoking (4 categories), BMI (in kg/m^2^), height (in m), alcohol consumption (in grams/day), physical activity (categorised score), dietary quality (Alternative Healthy Eating Index 2010, Chiuve et al., J Nutr, 2012), socioeconomic status score (at the local area level, ranging from 1 to 10), education score (ordinal variable ranging from 1: “primary school” to 8:“postgraduate degree”)

**Supplementary Table 5**. Association (Rate Ratios (RR), 95% confidence intervals (95% CI) between five first-generation methylation-based measures of aging (per one standard deviation) and cancer risk in the Melbourne Collaborative Cohort Study

|  |  |  | ***AA-Horvath*** | | | ***AA-Hannum*** | | | ***IEAA-Horvath*** | | | ***IEAA-Hannum*** | | | ***EEAA*** | | |
| --- | --- | --- | --- | --- | --- | --- | --- | --- | --- | --- | --- | --- | --- | --- | --- | --- | --- |
| **Cancer type** | **N cases** |  | **RR** | **95% CI** | **P** | **RR** | **95% CI** | **P** | **RR** | **95% CI** | **P** | **RR** | **95% CI** | **P** | **RR** | **95% CI** | **P** |
| **Colorectal cancer** | 814 | Model 1 | 1.11 | 1.00-1.24 | 0.05 | 1.11 | 0.99-1.23 | 0.06 | 1.10 | 0.99-1.23 | 0.08 | 1.06 | 0.95-1.17 | 0.32 | 1.10 | 0.99-1.23 | 0.07 |
|  |  | Model 2 | 1.10 | 0.99-1.23 | 0.09 | 1.10 | 0.99-1.23 | 0.08 | 1.08 | 0.97-1.21 | 0.16 | 1.04 | 0.94-1.16 | 0.44 | 1.10 | 0.99-1.23 | 0.09 |
| **Gastric cancer** | 166 | Model 1 | 0.90 | 0.71-1.13 | 0.36 | 1.07 | 0.85-1.34 | 0.58 | 1.00 | 0.80-1.26 | 0.98 | 1.16 | 0.92-1.45 | 0.21 | 1.06 | 0.85-1.32 | 0.61 |
|  |  | Model 2 | 0.94 | 0.73-1.21 | 0.64 | 1.14 | 0.89-1.48 | 0.30 | 1.04 | 0.81-1.34 | 0.75 | 1.24 | 0.96-1.59 | 0.10 | 1.12 | 0.87-1.44 | 0.38 |
| **Kidney cancer** | 139 | Model 1 | 1.13 | 0.86-1.49 | 0.39 | 1.46 | 1.11-1.92 | 0.01 | 1.11 | 0.86-1.44 | 0.43 | 1.42 | 1.07-1.88 | 0.01 | 1.44 | 1.11-1.86 | 0.01 |
|  |  | Model 2 | 1.20 | 0.87-1.66 | 0.27 | 1.61 | 1.14-2.27 | 0.01 | 1.15 | 0.86-1.55 | 0.35 | 1.52 | 1.07-2.16 | 0.02 | 1.55 | 1.12-2.15 | 0.01 |
| **Lung cancer** | 327 | Model 1 | 0.99 | 0.85-1.16 | 0.91 | 1.09 | 0.93-1.29 | 0.29 | 0.96 | 0.83-1.11 | 0.60 | 1.04 | 0.89-1.21 | 0.64 | 1.14 | 0.97-1.35 | 0.12 |
|  |  | Model 2 | 1.03 | 0.87-1.22 | 0.71 | 1.09 | 0.92-1.30 | 0.31 | 1.00 | 0.85-1.17 | 0.99 | 1.04 | 0.89-1.23 | 0.60 | 1.15 | 0.96-1.37 | 0.12 |
| **Mature B-cell neoplasms** | 426 | Model 1 | 1.22 | 1.07-1.38 | <0.01 | 1.24 | 1.09-1.41 | <0.01 | 1.03 | 0.91-1.17 | 0.63 | 1.01 | 0.89-1.15 | 0.84 | 1.32 | 1.15-1.5 | <0.01 |
|  |  | Model 2 | 1.22 | 1.07-1.39 | <0.01 | 1.25 | 1.10-1.43 | <0.01 | 1.03 | 0.90-1.17 | 0.68 | 1.01 | 0.89-1.16 | 0.85 | 1.33 | 1.16-1.53 | <0.01 |
| **Prostate cancer** | 847 | Model 1 | 1.04 | 0.93-1.15 | 0.51 | 0.95 | 0.85-1.06 | 0.38 | 1.05 | 0.95-1.16 | 0.37 | 0.97 | 0.87-1.09 | 0.63 | 0.95 | 0.85-1.05 | 0.31 |
|  |  | Model 2 | 1.02 | 0.92-1.14 | 0.65 | 0.95 | 0.85-1.06 | 0.37 | 1.04 | 0.93-1.15 | 0.51 | 0.97 | 0.87-1.09 | 0.60 | 0.95 | 0.85-1.06 | 0.34 |
| **Urothelial cancers** | 404 | Model 1 | 1.02 | 0.88-1.18 | 0.80 | 1.06 | 0.92-1.22 | 0.45 | 1.01 | 0.88-1.16 | 0.90 | 1.00 | 0.87-1.15 | 1 | 1.09 | 0.95-1.26 | 0.23 |
|  |  | Model 2 | 1.03 | 0.89-1.20 | 0.66 | 1.05 | 0.90-1.22 | 0.56 | 1.03 | 0.89-1.19 | 0.74 | 1.00 | 0.87-1.17 | 0.95 | 1.07 | 0.92-1.25 | 0.39 |
| **All types** | 3,086 | Model 1 | 1.07 | 1.02-1.13 | 0.01 | 1.09 | 1.04-1.15 | <0.01 | 1.04 | 0.99-1.10 | 0.13 | 1.04 | 0.98-1.09 | 0.19 | 1.11 | 1.05-1.17 | <0.01 |
|  |  | Model 2 | 1.07 | 1.01-1.13 | 0.01 | 1.09 | 1.04-1.15 | <0.01 | 1.04 | 0.98-1.09 | 0.19 | 1.03 | 0.98-1.09 | 0.24 | 1.11 | 1.05-1.17 | <0.01 |

**Model 1**: No adjustment other than that provided by the matching variables (age, sex, country of birth (Australia / Northern Europe / Southern Europe), sample type (peripheral blood mononuclear cells / dried blood spots / buffy coats); lung cancer study: additional matching for smoking status (never; former, quitting <10 years; former, quitting ≥10 years; current, smoking <15 cigarettes/day; current smoking ≥15 cigarettes/day)

**Model 2**: Additional adjustment for smoking (current / former / never), smoking pack-years, age at starting smoking (4 categories), time since quitting smoking (4 categories), BMI (in kg/m^2^), height (in m), alcohol consumption (in grams/day), physical activity (categorised score), dietary quality (Alternative Healthy Eating Index 2010, Chiuve et al., J Nutr, 2012), socioeconomic status score (at the local area level, ranging from 1 to 10), education score (ordinal variable ranging from 1: “primary school” to 8:“postgraduate degree”)

AA-Horvath: Age acceleration from the Horvath clock; AA-Hannum; Age acceleration from the Hannum clock; IEAA-Horvath: intrinsic epigenetic age acceleration from the Horvath clock; IEAA-Hannum: intrinsic epigenetic age acceleration from the Horvath clock; EEAA: enhanced Hannum age acceleration
